## Supplementary material for "Biomarkers of ageing and lung function trajectory: a multi-omics study in a UK birth cohort": Online Supplement – Supplementary Figures, Tables and Methods

**Authors**

Grenville Jack S, Yousefi Paul, Power Grace M, Richmond Rebecca C, Dodd James W

**Contents**

Page S2. Supplementary Figures

Page S3. Supplementary Tables

Page S14. Supplementary Methods

Page S17. Supplementary References

### Supplementary Figures

#### Figure S1. Participant flow for DNA methylation and proteomic exposures

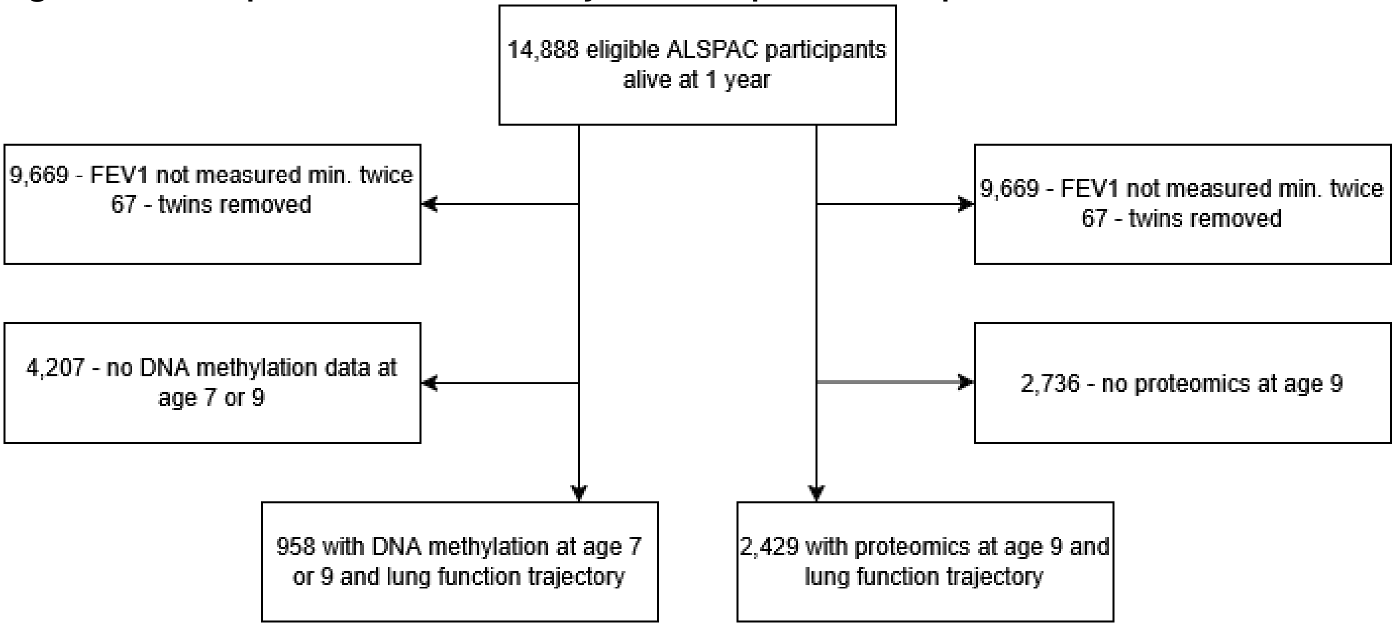

### Supplementary Tables

**Table S1. Characteristics of epigenetic clocks providing different measures of epigenetic age acceleration in this study**

| Clock | Generation | Training approach | Training sample | Target estimate |
|---|---|---|---|---|
| **Horvath**[1] | First-generation | DNA methylation at 353 CpGs selected using elastic net regression to predict chronological age | ~8,000 samples; 82 datasets; 51 tissue/cell types (mixed adult + paediatric) | Chronological age (multi-tissue 'pan-tissue' clock) |
| **Hannum**[2] | First-generation | DNA methylation at 71 CpGs in blood selected using elastic net regression to predict chronological age | n = 482; whole blood; adults (aged 19–101) | Chronological age (blood-specific) |
| **Wu**[3] | First-generation | DNA methylation at 111 CpGs in blood selected using elastic net regression to predict chronological age in children | n = 716 healthy children (9–212 months); blood; 11 cohorts | Chronological age in children (blood-specific) |
| **PhenoAge**[4] | Second-generation | Routine blood markers (albumin, creatinine, CRP, white cell counts) combined using penalized Cox regression into a mortality-risk score; DNA methylation at 513 CpGs then selected using elastic net regression to predict that score | n = 9,926 (NHANES III, mortality-risk score step); n = 456 (InCHIANTI, DNA methylation step) | Morbidity / mortality risk |
| **GrimAge (v1)**[5] | Second-generation | DNA methylation (1,030 CpGs across surrogates) selected using elastic net regression to predict plasma protein levels and smoking pack-years; these surrogates combined using elastic net Cox regression to predict time-to-death | n = 2,356 (Framingham Heart Study Offspring Cohort; 1,731 training / 625 test) | Mortality risk / lifespan |
| **DunedinPACE**[6] | Second-generation | DNA methylation at 173 CpGs selected using elastic net regression to predict the rate of decline across 19 | n = 1,037; single birth cohort (Dunedin Study) | Pace of biological ageing (rate |

| Clock | Generation | Training approach | Training sample | Target estimate |
|---|---|---|---|---|
| | (pace-of-aging) | organ-system biomarkers, measured repeatedly over ~20 years | followed longitudinally | per calendar year) |
| **CausAge**[7] | Third-generation (causality-enriched) | DNA methylation at CpGs causally linked to ageing traits (via Mendelian randomisation), selected using a causality-enriched elastic net regression to predict chronological age (586 CpGs) | n = 2,664 blood samples; adults | Composite causal epigenetic age (586 causally-implicated CpGs) |
| **DamAge**[7] | Third-generation (causality-enriched) | Same causally-linked CpG pool as CausAge, split by direction of effect; DamAge includes 1,090 deleterious CpGs | n = 2,664; same MR-derived CpG pool as CausAge | Accumulated damaging methylation changes; associates with mortality and adverse outcomes |
| **AdaptAge**[7] | Third-generation (causality-enriched) | Same causally-linked CpG pool as CausAge, split by direction of effect; AdaptAge includes 1,000 protective CpGs | n = 2,664; same MR-derived CpG pool as CausAge | Accumulated adaptive methylation changes; associates with protective outcomes |

**Table S2. Senescence-associated plasma proteins: relevance to cellular senescence, airway inflammation, lung function impairment and COPD**

| Protein | Relevance to cellular senescence | Relevance to airway inflammation, lung function or COPD | Refs. |
|---|---|---|---|
| **CCL11** | Induces fibroblast senescence via DNA damage response (p53, γH2AX); triggers SASP secretion (IL6, IL8) | Eosinophil chemoattractant in asthma; promotes fibroblast proliferation and collagen deposition (airway remodelling) | [8, 9] |
| **CXCL1** | NF-κB-driven SASP component | Epithelial-derived neutrophil/monocyte chemoattractant; elevated in COPD sputum | [8, 10, 11] |
| **CXCL5** | SASP chemokine | Neutrophil chemoattractant elevated in airways during COPD exacerbations | [8, 10, 12] |
| **CXCL10 (IP-10)** | IFN-driven SASP chemokine; recruits CD8+ T cells | CXCR3 ligand; elevated in COPD airways and correlates with radiological emphysema | [8, 13, 14] |
| **CXCL11** | Co-regulated with CXCL10, secretion in cellular senescence recruits Th1 and NK cells | CXCR3 ligand; CXCR3 knockout shown to reduce tobacco-smoke induced lung inflammation in animal models | [8, 15] |
| **IL-1α** | Upstream master regulator/initiator of the SASP via NF-κB | Serum levels correlate with airflow obstruction in asthma. Promotes lung inflammation, fibroblast activation and fibrosis in COPD | [8, 16, 17] |
| **IL-6** | Archetypal SASP effector; sustains senescence via paracrine autocrine loops | Serum level elevated in COPD and predicts exacerbations | [8, 18-20] |
| **IL-8 (CXCL8)** | Core SASP cytokine | Major neutrophil chemoattractant; elevated in COPD sputum | [8, 10, 21] |
| **IL-33** | Epithelial alarmin released on stress/damage; feeds IL-6/CXCL8 SASP loop | Drives eosinophil/Th2 inflammation in COPD. Blockade shown to be effective in reducing COPD exacerbations | [22, 23] |
| **LAP-TGF-β1** | Reinforces senescence and myofibroblast persistence via a senescence-fibrogenic loop | Drives epithelial-mesenchymal transition and airway remodelling; increased in bronchial reticular basement membrane and blood in COPD | [24-26] |

| Protein | Relevance to cellular senescence | Relevance to airway inflammation, lung function or COPD | Refs. |
|---|---|---|---|
| **MCP-1 (CCL2)** | SASP chemokine; recruits macrophages, reinforcing the senescent microenvironment | Elevated in COPD sputum/BAL, blood levels higher in emphysema | [27, 28] |
| **MMP-1** | SASP protease; extracellular-matrix-degrading effector of senescent cells | Interstitial collagenase; directly implicated in emphysema via collagen degradation in lung parenchyma | [29] |
| **SIRT2** | p53-dependent senescence marker; increases in stress-, replicative-, and oncogene-induced senescence | Sirtuin family important to accelerated ageing biology implicated in COPD pathogenesis | [30, 31] |

*Abbreviations – SASP – senescence-associated secretory phenotype; COPD – chronic obstructive pulmonary disease.

**Table S3. Five class $FEV_1$ trajectory model - trajectory group sizes and mean posterior probability of trajectory class assignment**

| Trajectory class | n | % | Mean posterior probability |
|---|---|---|---|
| Persistently high | 23 | 0.4 | 0.84 |
| Above average | 737 | 14.1 | 0.78 |
| Average | 2891 | 55.3 | 0.78 |
| Below average | 1423 | 27.2 | 0.75 |
| Persistently low | 158 | 3.0 | 0.78 |

*Reproduced from Grenville et al.[32]

**Table S4. Descriptive statistics for plasma protein concentrations at age 9**

| Plasma protein | Mean (NPX - $log^2$ scale) | Median (NPX - $log^2$ scale) | SD |
|---|---|---|---|
| CCL11 | 9.37 | 9.4 | 0.29 |
| CXCL1 | 9.35 | 9.3 | 0.68 |
| CXCL5 | 11.04 | 11.03 | 1.14 |
| CXCL10 | 9.34 | 9.17 | 0.89 |
| CXCL11 | 11.25 | 11.2 | 0.75 |
| IL1-α | -1.45 | -1.54 | 0.45 |
| IL6 | 3.28 | 3.13 | 0.71 |
| IL8 | 4.65 | 4.62 | 0.48 |
| IL33 | 1.6 | 1.57 | 0.32 |
| LAPTGFbeta1 | 7.98 | 7.96 | 0.37 |
| MCP1 | 12.49 | 12.48 | 0.4 |

| | | | |
|---|---|---|---|
| MMP1 | 12.3 | 12.24 | 1.06 |
| SIRT2 | 3.68 | 3.52 | 0.82 |

*NPX – normalised protein expression on a log[2] scale – one unit increase represents a doubling in protein concentration

**Table S5. Descriptive statistics for chronological age and epigenetic age (ages 7 or 9) and statistics regarding margins of error in age estimation for epigenetic clocks**

| | Mean | Median | SD | MAE | RMSE | Bias |
|---|---|---|---|---|---|---|
| Chronological age | 7.71 | 7.42 | 0.77 | - | - | - |
| Horvath | 8.95 | 8.45 | 3.06 | 2.28 | 3.35 | 1.25 |
| Hannum | 8.96 | 9.03 | 4.88 | 4.14 | 5.20 | 1.25 |
| Wu | 1.59 | 1.61 | 0.50 | 6.12 | 6.18 | -6.12 |
| PhenoAge | -2.20 | -2.00 | 7.16 | 10.44 | 12.37 | -9.92 |
| GrimAge v1 | 12.38 | 12.28 | 3.08 | 4.84 | 5.63 | 4.66 |
| CausAge | 0.34 | 0.18 | 4.91 | 7.70 | 8.80 | -7.37 |

*All figures are in units of years. Abbreviations – SD – standard deviation, MAE – mean absolute error, RMSE – root mean squared error.*

**Table S6. Odds ratios for Persistently Low lung function trajectory compared to others per standard deviation increase in different measures of epigenetic age acceleration**

| Exposure | Odds ratio (95% CI; FDR) – Model 1 (n = 958) | Odds ratio (95% CI; FDR) - Model 2 (n=872) |
|---|---|---|
| IEAA | 0.92 (0.58,1.33; FDR=0.85) | 0.98 (0.60, 1.43; FDR=0.92) |
| EEAA | 1.41 (0.96, 2.07; FDR=0.27) | 1.41 (0.92, 2.15; FDR=0.39) |
| Wu EAA | 0.97 (0.66, 1.43; FDR=0.88) | 1.09 (0.71, 1.66; FDR=0.80) |
| PhenoAge EAA | 1.17 (0.79, 1.73; FDR=0.78) | 1.08 (0.71, 1.65; FDR=0.80) |
| GrimAge EAA | 1.04 (0.68, 1.57; FDR=0.88) | 1.12 (0.72, 1.72; FDR=0.80) |
| DunedinPACE | 1.40 (0.94, 2.10; FDR=0.30) | 1.42 (0.92, 2.22; FDR=0.39) |
| DamAge EAA | 1.42 (0.95, 2.11; FDR=0.88) | 1.39 (0.91, 2.13; FDR=0.39) |
| AdaptAge EAA | 0.81 (0.54, 1.20; FDR=0.66) | 0.81 (0.53, 1.23; FDR=0.75) |
| CausAge EAA | 1.06 (0.72, 1.55; FDR=0.88) | 1.11 (0.73, 1.69; FDR=0.80) |

*Model 1 adjusted for age at blood sampling and sex. Model 2 adjusted for age at blood sampling, sex, maternal education ≤'O' level, prenatal maternal smoking, birth weight and BMI at age 8. Abbreviations:, FDR – false discovery rate, IEAA – intrinsic epigenetic age acceleration (Horvath clock), EEAA – extrinsic epigenetic age acceleration (Hannum clock), EAA – epigenetic age acceleration

**Table S7. Odds ratios for Below Average or Persistently Low lung function trajectories compared to others per standard deviation increase in different measures of epigenetic age acceleration**

| Exposure | Odds ratio (95% CI; FDR) – Model 1 (n = 958) | Odds ratio (95% CI; FDR) - Model 2 (n=872) |
|---|---|---|
| IEAA | 1.00 (0.87, 1.15; FDR=0.99) | 1.04 (0.90, 1.20; FDR=0.75) |
| EEAA | 1.02 (0.89, 1.17; FDR=0.99) | 1.03 (0.89, 1.19; FDR=0.80) |
| Wu EAA | 1.13 (0.98, 1.30; FDR=0.42) | 1.17 (1.01, 1.35; FDR=0.36) |
| PhenoAge EAA | 0.99 (0.86, 1.15; FDR=0.99) | 0.96 (0.83, 1.11; FDR=0.75) |
| GrimAge EAA | 1.14 (0.98, 1.32; FDR=0.42) | 1.14 (0.97, 1.33; FDR=0.47) |
| DunedinPACE | 1.07 (0.93, 1.24; FDR=0.79) | 1.08 (0.92, 1.25; FDR=0.72) |
| DamAge EAA | 1.11 (0.97; 1.28; FDR=0.43) | 1.10 (0.95, 1.28; FDR=0.56) |

| | | |
|---|---|---|
| AdaptAge EAA | 1.00 (0.87; 1.15; FDR=0.99) | 1.00 (0.86, 1.15; FDR=0.98) |
| CausAge EAA | 1.03 (0.90; 1.19; FDR=0.99) | 1.07 (0.92, 1.24; FDR=0.72) |

*Model 1 adjusted for age at blood sampling and sex. Model 2 adjusted for age at blood sampling, sex, maternal education ≤'O' level, prenatal maternal smoking, birth weight and BMI at age 8. Abbreviations: LCL – lower confidence limit, UCL – upper confidence limit, FDR – false discovery rate, IEAA – intrinsic epigenetic age acceleration (Horvath clock), EEAA – extrinsic epigenetic age acceleration (Hannum clock), EAA – epigenetic age acceleration

**Table S8. Odds ratios for Persistently Low lung function trajectory compared to others per standard deviation increase in each proteomic exposure**

| Exposure | Odds ratio (95% CI; FDR) – Model 1 (n = 2429) | Odds ratio (95% CI; FDR) - Model 2 (n = 2000) |
|---|---|---|
| CCL11 | 1.04 (0.80, 1.37; FDR=0.85) | 1.25 (0.92, 1.70; FDR=0.41) |
| CXCL1 | 1.15 (0.90, 1.47; FDR=0.48) | 1.11 (0.84, 1.44; FDR=0.65) |
| CXCL5 | 1.09 (0.84, 1.41; FDR=0.67) | 0.98 (0.75, 1.29; FDR=0.89) |
| CXCL10 | **1.29 (1.03, 1.57; FDR=0.08)*** | 1.24 (0.96, 1.54; FDR=0.33) |
| CXCL11 | 1.17 (0.91, 1.49; FDR=0.48) | 1.14 (0.86, 1.48; FDR=0.65) |
| IL1α | 0.88 (0.62, 1.15; FDR=0.62) | 0.87 (0.59, 1.15; FDR=0.65) |
| IL6 | **1.29 (1.03, 1.57; FDR=0.08)*** | 1.31 (1.03, 1.61; FDR=0.12) |
| IL8 | 1.12 (0.88, 1.41; FDR=0.55) | 1.10 (0.84, 1.44; FDR=0.65) |
| IL33 | **1.23 (1.04, 1.44; FDR=0.08)*** | 1.26 (1.05, 1.51; FDR=0.10) |
| LAPTGFβ1 | 1.06 (0.82, 1.36; FDR=0.75) | 1.09 (0.83, 1.42; FDR=0.65) |
| MCP1 | 1.24 (0.97, 1.57; FDR=0.27) | 1.22 (0.93, 1.57; FDR=0.41) |
| MMP1 | 1.17 (0.91, 1.50; FDR=0.48) | 1.09 (0.83, 1.43; FDR=0.65) |
| SIRT2 | 1.01 (0.76, 1.28; FDR=0.96) | 0.97 (0.71, 1.26; FDR=0.89) |

*Model 1 adjusted for age at blood sampling and sex. Model 2 adjusted for age at blood sampling, sex, maternal education ≤'O' level, prenatal maternal smoking, birth weight and BMI at age 8. Estimates represent increase in the odds of membership of the Persistently Low trajectory group per standard deviation increase in the plasma concentration of the respective proteomic exposure. Associations with significance robust to adjustment for multiple testing (FDR <0.10) are emboldened. Abbreviations: LCL – lower confidence limit, UCL – upper confidence limit, FDR – false discovery rate.

**Table S9. Odds ratios for Below Average or Persistently Low lung function trajectories compared to others per standard deviation increase in each proteomic exposure**

| Exposure | Odds ratio (95% CI; FDR) – Model 1 (n = 2429) | Odds ratio (95% CI; FDR) - Model 2 (n = 2000) |
|---|---|---|
| CCL11 | 0.97 (0.89, 1.07; FDR=0.68) | 0.97 (0.87, 1.08; FDR=0.70) |
| CXCL1 | 1.05 (0.96, 1.15; FDR=0.59) | 1.02 (0.93, 1.13; FDR=0.70) |
| CXCL5 | 1.09 (1.00, 1.19; FDR=0.59) | 1.03 (0.94, 1.14; FDR=0.70) |
| CXCL10 | 1.07 (0.98, 1.16; FDR=0.59) | 1.09 (0.99, 1.19; FDR=0.56) |
| CXCL11 | 1.02 (0.93, 1.12; FDR=0.68) | 1.03 (0.93, 1.14; FDR=0.70) |
| IL1α | 1.06 (0.97, 1.15; FDR=0.59) | 1.07 (0.98, 1.17; FDR=0.56) |
| IL6 | 1.03 (0.94, 1.12; FDR=0.68) | 1.07 (0.97, 1.18; FDR=0.56) |
| IL8 | 1.03 (0.94, 1.12; FDR=0.68) | 1.04 (0.95, 1.15; FDR=0.63) |
| IL33 | 1.03 (0.94, 1.12; FDR=0.68) | 1.05 (0.94, 1.15; FDR=0.63) |
| LAPTGFβ1 | 1.05 (0.96, 1.15; FDR=0.59) | 1.06 (0.97, 1.17; FDR=0.56) |
| MCP1 | 1.03 (0.94, 1.12; FDR=0.68) | 1.05 (0.95, 1.15; FDR=0.63) |
| MMP1 | 1.02 (0.93, 1.11; FDR=0.73) | 1.01 (0.91, 1.11; FDR=0.92) |
| SIRT2 | 1.05 (0.96, 1.15; FDR=0.59) | 1.06 (0.96, 1.17; FDR=0.56) |

*Model 1 adjusted for age at blood sampling and sex. Model 2 adjusted for age at blood sampling, sex, maternal education ≤'O' level, prenatal maternal smoking, birth weight and BMI at age 8. Estimates represent increase in the odds of membership of the Below Average or Persistently Low trajectory group, per standard deviation increase in the plasma concentration of the respective proteomic exposure. Results robust to adjustment for multiple testing (FDR <0.10) are emboldened. Abbreviations: LCL – lower confidence limit, UCL – upper confidence limit, FDR – false discovery rate.

### Table S10. Genetic instruments used in Mendelian randomisation analyses with associated first-stage statistics

| Exposure | Instrument | N | $R^2$ exposure (%) | F statistic | p value |
|---|---|---|---|---|---|
| CXCL10 | rs11548618 | 2,014 | 10.6 | 236.5 | $1.57 \times 10^{-50}$ |
| | PGRS (4 SNPs) | 2,014 | 3.3 | 68.4 | $2.37 \times 10^{-16}$ |
| IL6 (O-link) | rs4129267 | 2,014 | 1.7 | 33.8 | $6.99 \times 10^{-9}$ |
| | PGRS (7 SNPs) | 2,014 | 1.8 | 37.5 | $1.09 \times 10^{-9}$ |
| Log IL6 (pg/ml, ELISA) | rs4129267 | 2,635 | 2.3 | 63.1 | $2.93 \times 10^{-15}$ |
| | PGRS (7 SNPs) | 2,635 | 2.5 | 66.3 | $6.01 \times 10^{-16}$ |
| IL33 | rs185705622 | 2,014 | $3.26 \times 10^{-4}$ | 0.65 | 0.42 |
| $FEV_1$ | PGRS (554 SNPs) | 2,047 | 2.2 | 46.1 | $1.48 \times 10^{-11}$ |
| Asthma | PGRS (80 SNPs) | 2,085 | 1.0 | 20.0 | $8.35 \times 10^{-6}$ |

*Abbreviations: PGRS – polygenic risk score, SNP – single nucleotide polymorphism. P values correspond to the test of association between the genetic instrument and the exposure.

### Table S11. Results from two-stage residual inclusion Mendelian randomisation and logistic regression analyses of effect of change in plasma protein concentration on the odds of membership of the Persistently Low lung function trajectory

| Exposure | Method | OR | 95% LCL | 95% UCL | p | MDE 80% (OR) |
|---|---|---|---|---|---|---|
| CXCL10 | LR | 1.29 | 1.03 | 1.57 | 0.02 | - |
| | 2SRI MR | 1.18 | 0.58 | 2.37 | 0.65 | 2.72 |
| IL6 | LR | 1.29 | 1.03 | 1.57 | 0.02 | - |
| | 2SRI MR | 0.26 | 0.02 | 2.70 | 0.26 | 0.04 |

*Abbreviations: OR – odds ratio for Persistently Low lung function trajectory per standard deviation increase in plasma protein concentration, LCL – lower confidence limit, UCL – upper confidence limit, MDE 80% - minimum detectable OR at 80% power, LR – logistic regression, 2SRI MR – two-stage residual inclusion Mendelian randomisation. Where the corresponding odds ratio estimate is below 1, MDE 80% is enumerated as an odds ratio below 1.

### Table S12. Results from two-stage least squares Mendelian randomisation and ordinary least squares analyses of effect of change in plasma protein concentration on the time-weighted mean of $FEV_1$ ($FEV_1$ % AUC/time)

| Exposure | Method | β | 95% LCL | 95% UCL | p | MDE 80% | DWH p |
|---|---|---|---|---|---|---|---|
| CXCL10 | OLS | -0.56 | -1.04 | -0.08 | 0.02 | - | 0.92 |
| | 2SLS MR | -0.51 | -2.36 | 1.35 | 0.59 | 2.65 | |
| IL6 | OLS | -0.35 | -0.86 | 0.15 | 0.17 | - | 0.28 |
| | 2SLS MR | 0.17 | -3.75 | 4.09 | 0.93 | 5.61 | |

*Abbreviations: β – mean difference in $FEV_1$ % AUC/time per SD increase in plasma protein concentration, LCL – lower confidence limit, UCL – upper confidence limit, MDE 80% - minimum detectable mean difference in $FEV_1$ % AUC/time at 80% power, DWH P – p value for Durbin-Wu-Hauman test assessing for endogeneity in OLS estimate, OLS – ordinary least squares, 2SLS MR – two-stage least squares Mendelian randomisation

**Table S13. Proportion variance explained in exposure in forwards and reverse MR analyses with continuous exposures and outcomes**

| Exposure | Instrument | Outcome | $R^2$ exposure (%) | $R^2$ outcome (%) |
|---|---|---|---|---|
| CXCL10 | rs11548618 | $FEV_1$ % AUC/time | 11.4 | 0.8 |
| IL6 | rs4129267 | $FEV_1$ % AUC/time | 3.2 | 0.7 |
| $FEV_1$ | PGS (554 SNPs) | CXCL10 (age 9) | 2.2 | 0.3 |
| $FEV_1$ | PGS (554 SNPs) | CXCL10 (age 24) | 2.3 | 0.5 |
| $FEV_1$ | PGS (554 SNPs) | IL6 (age 9) | 2.2 | 0.1 |
| $FEV_1$ | PGS (554 SNPs) | IL6 (age 24) | 2.3 | 0.1 |
| $FEV_1$ | PGS (554 SNPs) | IL33 (age 9) | 2.2 | 0.1 |
| $FEV_1$ | PGS (554 SNPs) | IL33 (age 24) | 2.3 | 0.0 |

*Abbreviations: $FEV_1$ % AUC/time – time weighted mean of $FEV_1$ % predicted. PGRS – polygenic score, SNP – single nucleotide polymorphism.

**Table S14. Association of genetic instruments for plasma proteins in primary analyses with potential confounders of the relationship between proteins and longitudinal $FEV_1$**

| Instrument (exposure) | Potential confounder | β (95% CI; p) |
|---|---|---|
| rs11548618 (CXCL10) | Maternal education ≤'O' level | -0.05 (-0.14, 0.04; p=0.28) |
| | Prenatal maternal smoking | -0.02 (-0.07, 0.02; p=0.32) |
| | Birth weight | 0.00 (-0.08, 0.09; p=0.95) |
| | BMI at age 8 | -0.34 (-0.89, 0.20; p=0.21) |
| rs4129267 (IL6) | Maternal education ≤'O' level | 0.00 (-0.01, 0.01; p=0.99) |
| | Prenatal maternal smoking | 0.00 (0.00, 0.01; p=0.51) |
| | Birth weight | -0.01 (-0.02, 0.01; p=0.45) |
| | BMI at age 8 | 0.05 (-0.04, 0.15; p=0.26) |

*Abbreviations: β – mean difference in continuous variable, or mean difference in log odds of binary variable, per additional effect alelle, CI – confidence interval, BMI – body mass index

**Table S15. Results from two-stage residual inclusion Mendelian randomisation analyses incorporating different instrument-exposure pairings to estimate effects of plasma CXCL10 and IL6 on the odds of Persistently Low lung function trajectory**

| Exposure | Instrument | OR | 95% LCL | 95% UCL | p | MDE 80% (OR) |
|---|---|---|---|---|---|---|
| CXCL10 | PGS (4 SNPs) | 0.43 | 0.05 | 2.33 | 0.34 | 0.08 |
| IL6 (O-link) | PGS (7 SNPs) | 0.25 | 0.02 | 2.29 | 0.27 | 0.04 |
| Log IL6 (pg/ml - ELISA) | rs4129267 | 0.57 | 0.10 | 2.69 | 0.53 | 0.10 |
| Log IL6 (pg/ml - ELISA) | PGS (7 SNPs) | 0.54 | 0.10 | 2.42 | 0.49 | 0.10 |

*Abbreviations: OR – odds ratio for Persistently Low lung function trajectory per standard deviation increase in plasma protein concentration, LCL – lower confidence limit, UCL – upper confidence limit, MDE 80% - minimum detectable OR at 80% power, ELISA - Enzyme-Linked Immunosorbent Assay, PGS – polygenic score. Where the corresponding odds ratio estimate is below 1, MDE 80% is enumerated as an odds ratio below 1.

**Table S16. Results from two-stage least squares Mendelian randomisation analyses incorporating different instrument-exposure pairings to estimate effects of plasma CXCL10 and IL6 on the time-weighted mean of $FEV_1$ ($FEV_1$ % AUC/time)**

| Exposure | Instrument | β | 95% LCL | 95% UCL | p | MDE 80% |
|---|---|---|---|---|---|---|
| CXCL10 | PGS (4 SNPs) | 0.71 | -2.20 | 3.63 | 0.63 | 4.17 |
| IL6 (O-link) | PGS (7 SNPs) | 0.01 | -3.69 | 3.72 | 0.99 | 5.30 |
| Log IL6 (pg/ml - ELISA) | rs4129267 | 0.24 | -2.64 | 3.11 | 0.87 | 4.11 |
| Log IL6 (pg/ml - ELISA) | PGS (7 SNPs) | 0.01 | -2.77 | 2.80 | 0.99 | 3.98 |

*Abbreviations: β – mean difference in $FEV_1$ % AUC/time per standard deviation increase in genetically predicted IL6 concentration, LCL – lower confidence limit, UCL – upper confidence limit, MDE 80% - minimum detectable OR at 80% power, ELISA - Enzyme-Linked Immunosorbent Assay, PGS – polygenic score

**Table S17. Results from two-stages least squares Mendelian randomisation and ordinary least squares analyses of the effect of childhood $FEV_1$ on plasma protein concentrations at ages 9 and 24**

| Outcome | Method | β | 95% LCL | 95% UCL | p | MDE 80% |
|---|---|---|---|---|---|---|
| CXCL10 (age 9) | OLS | -0.04 | -0.08 | -0.001 | 0.04 | |
| | 2SLS MR | -0.34 | -0.65 | -0.03 | 0.03 | 0.44 |
| CXCL10 (age 24) | OLS | -0.05 | -0.08 | -0.01 | 0.02 | |
| | 2SLS MR | -0.38 | -0.67 | -0.08 | 0.01 | 0.42 |
| IL6 (age 9) | OLS | -0.02 | -0.05 | 0.01 | 0.25 | |
| | 2SLS MR | -0.16 | -0.39 | 0.07 | 0.18 | 0.33 |
| IL6 (age 24) | OLS | -0.04 | -0.08 | 0.00 | 0.05 | |
| | 2SLS MR | -0.17 | -0.44 | 0.10 | 0.22 | 0.39 |
| IL33 (age 9) | OLS | 0.001 | -0.02 | 0.02 | 0.94 | |
| | 2SLS MR | -0.09 | -0.18 | 0.01 | 0.08 | 0.13 |
| IL33 (age 24) | OLS | -0.02 | -0.03 | -0.002 | 0.03 | |
| | 2SLS MR | -0.002 | -0.10 | 0.09 | 0.97 | 0.14 |

*Abbreviations: β – mean difference in plasma protein concentration per standard deviation increase in $FEV_1$, LCL – lower confidence limit, UCL – upper confidence limit, MDE 80% - minimum detectable OR at 80% power, OLS – ordinary least squares, 2SLS MR – two-stage least squares Mendelian randomisation

**Table S18. Results from two-stages least squares Mendelian randomisation and ordinary least squares analyses of the effect of childhood asthma diagnosis on plasma protein concentrations at ages 9 and 24**

| Outcome | Method | β | 95% LCL | 95% UCL | p | MDE 80% |
|---|---|---|---|---|---|---|
| CXCL10 (age 9) | OLS | 0.13 | -0.08 | 0.11 | 0.79 | |
| | 2SLS MR | -0.69 | -1.76 | 0.38 | 0.21 | 1.53 |
| CXCL10 (age 24) | OLS | 0.13 | 0.03 | 0.23 | 0.05 | |
| | 2SLS MR | 0.21 | -0.50 | 0.92 | 0.56 | 1.01 |
| IL6 (age 9) | OLS | 0.06 | -0.01 | 0.14 | 0.10 | |
| | 2SLS MR | -0.65 | -1.54 | 0.24 | 0.15 | 1.27 |
| IL6 (age 24) | OLS | 0.07 | -0.02 | 0.16 | 0.12 | |
| | 2SLS MR | 0.15 | -0.57 | 0.87 | 0.69 | 1.03 |
| IL33 (age 9) | OLS | 0.01 | -0.03 | 0.05 | 0.62 | |
| | 2SLS MR | 0.05 | -0.32 | 0.43 | 0.78 | 0.53 |
| IL33 (age 24) | OLS | -0.02 | -0.04 | 0.01 | 0.16 | |
| | 2SLS MR | -0.27 | -0.53 | -0.01 | 0.04 | 0.37 |

*Abbreviations: β – mean difference in standardised plasma CXCL10 concentration per standard deviation increase in genetic liability to childhood asthma, LCL – lower confidence limit, UCL – upper confidence limit, MDE 80% - minimum detectable OR at 80% power, OLS – ordinary least squares, 2SLS MR – two-stage least squares Mendelian randomisation

**Table S19. Association of $FEV_1$ polygenic score with potential confounders of the relationship between childhood $FEV_1$ and plasma proteins**

| Potential confounder | β (95% CI; p) |
|---|---|
| Maternal education ≤16 years | 0.00 (-0.01, 0.01; p=0.45) |
| Prenatal maternal smoking | 0.00 (0.00, 0.00; p=0.93) |
| Birth weight | 0.01 (0.00, 0.02; p=0.16) |
| BMI at age 8 | -0.10 (-0.17, -0.04; p=0.00) |

*Abbreviations: β – mean difference in continuous variable, or mean difference in log odds of binary variable, per standard deviation increase in polygenic risk score for $FEV_1$, CI – confidence interval, BMI – body mass index

**Table S20. One-sample and two-sample Mendelian randomisation estimates for effect of genetically predicted childhood $FEV_1$ on plasma proteins**

| Outcome | MR estimate | β (95% CI; p) | p for Cochrane's Q |
|---|---|---|---|
| CXCL10 (age 9) | 2SLS with PGS for $FEV_1$ | -0.34 (-0.65, -0.03; p=0.03) | |
| | Inverse-variance weighted | -0.04 (-0.19, 0.10; p=0.56) | 0.12 |
| | Weighted median | 0.00 (-0.22, 0.23; p=0.97) | |
| | Weighted mode | 0.13 (-0.56, 0.81; p=0.72) | |
| | Egger intercept | 0.00 (0.00, 0.01; p=0.24) | |
| CXCL10 (age 24) | 2SLS with PGS for $FEV_1$ | -0.38 (-0.67, -0.08, p=0.01) | |
| | Inverse-variance weighted | -0.12 (-0.26, 0.01; p=0.07) | 0.35 |
| | Weighted median | -0.08 (-0.29, 0.14; p=0.48) | |
| | Weighted mode | 0.16 (-0.49, 0.81; p=0.63) | |
| | Egger intercept | 0.00 (0.00, 0.01; p=0.45) | |
| IL6 (age 9) | 2SLS with PGS for $FEV_1$ | -0.16 (-0.39, 0.07; p=0.18) | |
| | Inverse-variance weighted | -0.05 (-0.16, 0.07; p=0.40) | 0.03 |
| | Weighted median | 0.00 (-0.18, 0.18; p=0.98) | |
| | Weighted mode | 0.13 (-0.37, 0.62; p=0.62) | |
| | Egger intercept | 0.00 (0.00, 0.01; p=0.30) | |
| IL6 (age 24) | 2SLS with PGS for $FEV_1$ | -0.17 (-0.44, 0.10; p=0.22) | |
| | Inverse-variance weighted | -0.12 (-0.25, 0.01; p=0.07) | 0.55 |
| | Weighted median | -0.09 (-0.30, 0.12; p=0.38) | |
| | Weighted mode | 0.19 (-0.40, 0.79; p=0.52) | |
| | Egger intercept | 0.00 (-0.01, 0.01; p=0.66) | |
| IL33 (age 9) | 2SLS with PGS for $FEV_1$ | -0.09 (-0.18, 0.01; p=0.08) | |
| | Inverse-variance weighted | -0.04 (-0.09, 0.01; p=0.16) | 0.39 |
| | Weighted median | -0.06 (-0.14, 0.02; p=0.16) | |
| | Weighted mode | -0.12 (-0.34, 0.10; p=0.28) | |
| | Egger intercept | 0.00 (0.00, 0.00; p=0.13) | |
| IL33 (age 24) | 2SLS with PGS for $FEV_1$ | 0.00 (-0.10, 0.09; p=0.97) | |
| | Inverse-variance weighted | -0.01 (-0.06, 0.04; p=0.73) | 0.65 |
| | Weighted median | -0.03 (-0.11, 0.05; p=0.45) | |
| | Weighted mode | -0.01 (-0.21, 0.18; p=0.90) | |
| | Egger intercept | -0.04 (-0.09, 0.01; p=0.16) | |

*Abbreviations: β – mean difference in standardised plasma protein concentration per standard deviation increase in genetically predicted childhood $FEV_1$, CI – confidence interval, 2SLS – two-stage least squares, PGS – polygenic score

### Supplementary Methods

#### Calculation of specific measures of epigenetic age acceleration

Intrinsic epigenetic age acceleration (IEAA) was calculated with inclusion of peripheral blood cell counts as covariates in the regression model for Horvath age acceleration.[33] Extrinsic Epigenetic Age Acceleration (EEAA) was calculated from the Hannum DNAm age estimate by applying published weighting coefficients to estimated peripheral cell counts.[33, 34] DunedinPACE was derived from longitudinal associations of DNA methylation with phenotypic ageing traits and estimates the pace of biological aging; values are centred on 1, with values under 1 indicating slower aging and over 1 faster aging.[6]

#### Genotype data and quality control procedures

Genotyping was performed with the Illumina HumanHap550 quad array. Samples were removed during quality control in case of sex discrepancies, minimal or excessive heterozygosity, high missingness (>3%) or insufficient sample replication (IBD <0.8). Individuals of European ancestry were retained after multidimensional scaling analysis and comparison with reference panels to assess for population stratification.

Single nucleotide polymorphisms (SNPs) were removed if they did not meet the following quality control thresholds: missing call rate >1%, minor allele frequency (MAF) <0.01, or Hardy–Weinberg equilibrium (HWE) P $<1^{-6}$. Related individuals were included. Imputation of SNPs in the remaining dataset was performed to the TOPMed reference panel[35] (Genome build GrCh38; phasing with Eagle v2.4; imputation with Minimac v1.0.2), with an imputation quality ($R^2$) threshold of 0.3. 455,150 autosomal SNPs were included in the final dataset. SNPs extracted for MR were required to have an imputation $R^2$ of ≥0.8.

#### Details of genome-wide association studies and specifications to identify genetic instruments

Genetic instruments for proteomic exposures were genome-wide significant ($p < 5 \times 10^{8}$) single nucleotide polymorphisms (SNPs) obtained from a GWAS of 2,923 plasma proteins in 54,219 UK Biobank participants, prioritising genome-wide significant cis- protein quantitative trait loci (pQTLs) within 10 Mb of the encoding gene for protein or receptor.[36] In primary analyses, the strongest available genome-wide significant cis-pQTL was used as a single-SNP instrument for the corresponding protein, without application of a threshold for minor allele frequency in order to retain a cis-instrument for proteins where the only available cis-pQTL was rare. Sensitivity analyses used weighted polygenic scores (PGS) incorporating all cis- and trans-pQTLs from the same GWAS, with weights determined by the original GWAS beta coefficients. Genomic positions (GrCh38) and weights for all pQTLs are provided in Supplementary Files 1-2.

For reverse MR, the instrument for childhood $FEV_1$ was a weighted PGS comprising genome-wide significant, linkage disequilibrium (LD) independent ($R^2$ <0.01, 10Mb window) SNPs from 475,645 European ancestry participants in the largest available GWAS meta-analysis (mean age 54.4 years).[37]

Sample overlap with ALSPAC was 1.02% (4,857 participants). The genetic instrument for childhood asthma was a weighted PGS constructed from genome-wide significant lead SNPs for self-reported childhood-onset asthma (≤19 years) from a European-ancestry only meta-analysis of FinnGenn and UK Biobank cohorts (20,964 cases, 674,509 controls).[38] No sample overlap with ALSPAC was identified. PGS incorporated weights from the parent GWAS and SNPs were excluded if failing the following quality control thresholds: missingness >1%, MAF <1%, HWE P <1x10 $^{-6}$. Prior to PGS construction, SNPs for $FEV_1$ were clumped in ALSPAC ($r^2$ ≥0.01, window 10 Mb) to ensure independence. Genomic positions (GrCh38) and weights of all SNPs in PGS for $FEV_1$ and asthma are included in Supplementary Files 3 and 4.

**Statistical procedures for Mendelian randomisation analyses**

Two-stage residual inclusion (2SRI) Mendelian randomisation (MR) involves first-stage regression of the exposure on the genetic instrument, followed by second stage regression of outcome on genetically predicted exposure with inclusion of first-stage residuals to adjust for residual first-stage confounding. 95% confidence intervals were derived from HC3-robust standard errors.[39]

Two-stage least squares (2SLS) MR was performed for continuous outcomes ($FEV_1$ % AUC/time, CXCL10, IL6 and IL33), involving second stage regression of the outcome on genetically predicted values of the exposure.

2SLS estimates were compared with those from ordinary-least-squares (OLS) linear regression, adjusting for the same covariates. The Durbin-Wu-Hausman test was applied to assess whether the 2SLS and OLS estimates were statistically distinguishable, with p<0.05 indicating evidence of endogeneity in the OLS estimate.

**Instrumental variable assumptions**

An evaluation of instrument strength for each MR exposure through calculation of relevant F statistics supported an assessment of satisfaction of the first assumption, that genetic instruments must be robustly associated with the exposure. Steiger directionality testing was also applied to check that a greater proportion of variance was explained in exposure than outcome.

To assess the independence assumption, that genetic instruments should not be associated with confounders of the exposure-outcome relationship, appropriate regression models were fitted with each covariate from adjusted observational models as outcome, genetic instrument as exposure, and sex and genetic principal components as covariates.

A direct statistical assessment of the exclusion restriction assumption, that genetic variants must influence the outcome only through the exposure, was not possible for our primary single-SNP MR analyses on proteomic exposures because such tests rely on MR models with multiple SNPs. To minimise the potential for horizontal pleiotropy, we utilised cis-variants within 10Mb of the gene encoding for protein or receptor.

In reverse MR, in which the genetic instrument for $FEV_1$ was a PGRS with multiple SNPs, evaluation of pleiotropy involved application of two-sample MR methods utilising SNP-exposure associations from GWAS summary statistics and SNP-outcome associations from our own analyses in ALSPAC. Exposure and outcome summary statistics were harmonised to ensure consistent alignment of effect alleles. SNP effects were oriented to the same effect allele, with allele frequencies used to resolve strand ambiguity where possible. Palindromic variants with ambiguous allele frequencies were excluded. We compared estimates from inverse variance weighted (IVW), weighted median and weighted mode methods. While IVW assumes no directional pleiotropy, weighted median assumes that a majority of instruments act only through the exposure, and weighted mode assumes this is true of a plurality. We also assessed heterogeneity of SNP effect in IVW through Cochrane's Q and evaluated variation from zero in the MR Egger intercept, which suggests directional pleiotropy if present.

**Protocol**

The analysis plan was detailed in the ALSPAC data access proposal approved before data release.

**Statistical packages**

Global lung function initiative per cent of predicted lung function values for age, sex and height were calculated using the *rspiro* package in R 4.4.3. To maintain consistency with the previous approach in ALSPAC, latent profile analysis was performed using the *gllam* package in Stata 18.0 (StataCorp LLC, TX, USA). Genetic data extraction and weighted polygenic risk score calculation were conducted with *PLINK* 2.0 (version 2.00a build 68). All other analyses employed R 4.4.3; the *ivreg* (0.6.5) package was used for one-sample MR and two-sample analyses utilised *TwoSampleMR* (0.6.24). Epigenetic age acceleration was calculated using *meffonym*.[40]

**Supplementary References**

1. Horvath S. DNA methylation age of human tissues and cell types. Genome biology. 2013;14:1-20.
2. Hannum G, Guinney J, Zhao L, et al. Genome-wide methylation profiles reveal quantitative views of human aging rates. Molecular cell. 2013;49(2):359-67.
3. Wu X, Chen W, Lin F, et al. DNA methylation profile is a quantitative measure of biological aging in children. Aging (Albany NY). 2019;11(22):10031-51. 10.18632/aging.102399
4. Levine ME, Lu AT, Quach A, et al. An epigenetic biomarker of aging for lifespan and healthspan. Aging (Albany NY). 2018;10(4):573-91. 10.18632/aging.101414
5. Lu AT, Quach A, Wilson JG, et al. DNA methylation GrimAge strongly predicts lifespan and healthspan. Aging (Albany NY). 2019;11(2):303-27. 10.18632/aging.101684
6. Belsky DW, Caspi A, Corcoran DL, et al. DunedinPACE, a DNA methylation biomarker of the pace of aging. Elife. 2022;11. 10.7554/eLife.73420
7. Ying K, Liu H, Tarkhov AE, et al. Causality-enriched epigenetic age uncouples damage and adaptation. Nature Aging. 2024;4(2):231-46. 10.1038/s43587-023-00557-0
8. Wang B, Han J, Elisseeff JH, Demaria M. The senescence-associated secretory phenotype and its physiological and pathological implications. Nature Reviews Molecular Cell Biology. 2024;25(12):958-78. 10.1038/s41580-024-00727-x
9. Isgrò M, Bianchetti L, Marini MA, Bellini A, Schmidt M, Mattoli S. The C-C motif chemokine ligands CCL5, CCL11, and CCL24 induce the migration of circulating fibrocytes from patients with severe asthma. Mucosal Immunology. 2013;6(4):718-27. 10.1038/mi.2012.109
10. Henrot P, Prevel R, Berger P, Dupin I. Chemokines in COPD: From Implication to Therapeutic Use. Int J Mol Sci. 2019;20(11). 10.3390/ijms20112785
11. Inui T, Watanabe M, Nakamoto K, et al. Bronchial epithelial cells produce CXCL1 in response to LPS and TNFα: A potential role in the pathogenesis of COPD. Exp Lung Res. 2018;44(7):323-31. 10.1080/01902148.2018.1520936
12. Qiu Y, Zhu J, Bandi V, et al. Biopsy neutrophilia, neutrophil chemokine and receptor gene expression in severe exacerbations of chronic obstructive pulmonary disease. Am J Respir Crit Care Med. 2003;168(8):968-75. 10.1164/rccm.200208-794OC
13. Saetta M, Mariani M, Panina-Bordignon P, et al. Increased Expression of the Chemokine Receptor CXCR3 and Its Ligand CXCL10 in Peripheral Airways of Smokers with Chronic Obstructive Pulmonary Disease. American Journal of Respiratory and Critical Care Medicine. 2002;165(10):1404-9. 10.1164/rccm.2107139
14. Hao W, Li M, Pang Y, Du W, Huang X. Increased chemokines levels in patients with chronic obstructive pulmonary disease: correlation with quantitative computed tomography metrics. Br J Radiol. 2021;94(1118):20201030. 10.1259/bjr.20201030
15. Nie L, Xiang R, Zhou W, Lu B, Cheng D, Gao J. Attenuation of acute lung inflammation induced by cigarette smoke in CXCR3 knockout mice. Respir Res. 2008;9(1):82. 10.1186/1465-9921-9-82
16. Evans MD, Esnault S, Denlinger LC, Jarjour NN. Sputum cell IL-1 receptor expression level is a marker of airway neutrophilia and airflow obstruction in asthmatic patients. J Allergy Clin Immunol. 2018;142(2):415-23. 10.1016/j.jaci.2017.09.035
17. Osei ET, Noordhoek JA, Hackett TL, et al. Interleukin-1α drives the dysfunctional cross-talk of the airway epithelium and lung fibroblasts in COPD. Eur Respir J. 2016;48(2):359-69. 10.1183/13993003.01911-2015
18. Barnes PJ, Baker J, Donnelly LE. Cellular Senescence as a Mechanism and Target in Chronic Lung Diseases. Am J Respir Crit Care Med. 2019;200(5):556-64. 10.1164/rccm.201810-1975TR

19. van Durme YM, Lahousse L, Verhamme KM, et al. Mendelian randomization study of interleukin-6 in chronic obstructive pulmonary disease. Respiration. 2011;82(6):530-8. 10.1159/000332336
20. Huang H, Huang X, Zeng K, Deng F, Lin C, Huang W. Interleukin-6 is a Strong Predictor of the Frequency of COPD Exacerbation Within 1 Year. Int J Chron Obstruct Pulmon Dis. 2021;16:2945-51. 10.2147/copd.S332505
21. Yamamoto C, Yoneda T, Yoshikawa M, et al. Airway inflammation in COPD assessed by sputum levels of interleukin-8. Chest. 1997;112(2):505-10. 10.1378/chest.112.2.505
22. Tworek D, Majewski S, Szewczyk K, et al. The association between airway eosinophilic inflammation and IL-33 in stable non-atopic COPD. Respir Res. 2018;19(1):108. 10.1186/s12931-018-0807-y
23. Sciurba FC, Watz H, Bourdin A, et al. Tozorakimab to Prevent COPD Exacerbations. N Engl J Med. 2026. 10.1056/NEJMoa2606998
24. Ojiaku CA, Yoo EJ, Panettieri RA, Jr. Transforming Growth Factor β1 Function in Airway Remodeling and Hyperresponsiveness. The Missing Link? Am J Respir Cell Mol Biol. 2017;56(4):432-42. 10.1165/rcmb.2016-0307TR
25. Soltani A, Sohal SS, Reid D, Weston S, Wood-Baker R, Walters EH. Vessel-associated transforming growth factor-beta1 (TGF-β1) is increased in the bronchial reticular basement membrane in COPD and normal smokers. PLoS One. 2012;7(6):e39736. 10.1371/journal.pone.0039736
26. Mak JCW, Chan-Yeung MMW, Ho SP, et al. Elevated plasma TGF-β1 levels in patients with chronic obstructive pulmonary disease. Respiratory Medicine. 2009;103(7):1083-9. https://doi.org/10.1016/j.rmed.2009.01.005
27. Traves SL, Culpitt SV, Russell RE, Barnes PJ, Donnelly LE. Increased levels of the chemokines GROalpha and MCP-1 in sputum samples from patients with COPD. Thorax. 2002;57(7):590-5. 10.1136/thorax.57.7.590
28. Di Stefano A, Coccini T, Roda E, et al. Blood MCP-1 levels are increased in chronic obstructive pulmonary disease patients with prevalent emphysema. Int J Chron Obstruct Pulmon Dis. 2018;13:1691-700. 10.2147/copd.S159915
29. Christopoulou ME, Papakonstantinou E, Stolz D. Matrix Metalloproteinases in Chronic Obstructive Pulmonary Disease. Int J Mol Sci. 2023;24(4). 10.3390/ijms24043786
30. Anwar T, Khosla S, Ramakrishna G. Increased expression of SIRT2 is a novel marker of cellular senescence and is dependent on wild type p53 status. Cell Cycle. 2016;15(14):1883-97. 10.1080/15384101.2016.1189041
31. Barnes PJ. Chapter Nine - Senotherapy for lung diseases. In: Wong WSF, editor. Advances in Pharmacology. 98: Academic Press; 2023. p. 249-71.
32. Grenville JS, Richmond RC, Power GM, Yousefi PD, Dodd JW. Causal effects of early-life traits on lung function trajectory: a Mendelian randomization study. medRxiv. 2026:2026.10.27.26366597. 10.64898/2026.10.27.26366597
33. Chen BH, Marioni RE, Colicino E, et al. DNA methylation-based measures of biological age: meta-analysis predicting time to death. Aging (Albany NY). 2016;8(9):1844-65. 10.18632/aging.101020
34. Quach A, Levine ME, Tanaka T, et al. Epigenetic clock analysis of diet, exercise, education, and lifestyle factors. Aging (Albany NY). 2017;9(2):419-46. 10.18632/aging.101168
35. Taliun D, Harris DN, Kessler MD, et al. Sequencing of 53,831 diverse genomes from the NHLBI TOPMed Program. Nature. 2021;590(7845):290-9. 10.1038/s41586-021-03205-y
36. Sun BB, Chiou J, Traylor M, et al. Plasma proteomic associations with genetics and health in the UK Biobank. Nature. 2023;622(7982):329-38. 10.1038/s41586-023-06592-6

37. Shrine N, Izquierdo AG, Chen J, et al. Multi-ancestry genome-wide association analyses improve resolution of genes and pathways influencing lung function and chronic obstructive pulmonary disease risk. Nature Genetics. 2023;55(3):410-22. 10.1038/s41588-023-01314-0
38. Tsuo K, Zhou W, Wang Y, et al. Multi-ancestry meta-analysis of asthma identifies novel associations and highlights the value of increased power and diversity. Cell Genom. 2022;2(12):100212. 10.1016/j.xgen.2022.100212
39. MacKinnon JG, White H. Some heteroskedasticity-consistent covariance matrix estimators with improved finite sample properties. Journal of Econometrics. 1985;29(3):305-25. https://doi.org/10.1016/0304-4076(85)90158-7
40. Suderman MaY, P. DNA methylation indices of exposure and phenotype (meffonym) [Internet]. 2026. Available from: https://github.com/perishky/meffonym (accessed 02/10/2026).
